# Developing Catchment Area Data Dashboards for Cancer Centers: A Stakeholder- engaged Approach

**DOI:** 10.1101/2024.07.05.24309999

**Authors:** Kalyani Sonawane, Ketki N Borse, Melanie Jefferson, Haluk Damgacioglu, Matthew J Carpenter, John L. Pearce, Besim Ogretmen, Sophie Paczesny, John P. O’Bryan, Jihad S. Obeid, Marvella E Ford, Ashish A Deshmukh

## Abstract

**Background:** Data dashboards that can communicate complex and diverse catchment area data effectively can transform cancer prevention and care delivery and strengthen community engagement efforts. Engaging stakeholders in data dashboard development, by seeking their inputs and collecting feedback, has the potential to maximize user-centeredness.

**Objective:** To describe a systematic, stakeholder-driven, and theory-based approach for developing catchment area data visualization tools for cancer centers.

**Methods:** Cancer-relevant catchment area data were identified from national- and state-level data sources (including cancer registries, national surveys, and administrative claims databases). A prototype tool for data visualization was designed, developed, and tested based on the *OPT-In* [*O*rganize, *P*lan, *T*est, *In*tegrate] framework. A working group of multi-disciplinary experts collected stakeholder feedback through formative assessment to understand data and design preferences. Thematic areas, data elements, and the composition and placement of data visuals in the prototype were identified and refined by working group members. Visualizations were rendered in Tableau^©^ and embedded in a public-facing website. A mixed-method approach was used to assess the understandability and actionability of the tool and to collect open-ended feedback that informed action items for improvisation.

**Results:** We developed a visualization dashboard that illustrates cancer incidence and mortality, risk factor prevalence, healthcare access, and social determinants of health for the Hollings Cancer Center catchment area. Color-coded maps, time-series plots, and graphs illustrate these catchment area data. A total of 21 participants representing key stakeholders [general audience (n=4), community advisory board members and other representatives (n=7), and researchers (n=10)] were identified. The understandability and actionability scores exceeded the minimum (80%) threshold. Stakeholders’ feedback confirmed that the tool is effective in communicating cancer data and is useful for education and advocacy. Themes that emerged from qualitative data suggest that additional changes to the tool such as a warm color palette, data source transparency, and the addition of analytical features (data overlaying and area-resolution selection) would further enhance the tool. Integration of communication efforts and messages within a broader context is in progress.

**Discussion:** A catchment area data resource developed through a systematic, stakeholder-driven, and theory-based approach can meet (and surpass) benchmarks for understandability and actionability, and lead to an overall positive response from stakeholders. Creating channels for advocacy and forming community partnerships will be the next step necessary to promote policies and programs for improving cancer outcomes in the catchment areas.

## INTRODUCTION

The idea of ‘cancer centers’ was first mentioned in the National Cancer Act of 1937.^1, 2^ About four decades later, the National Cancer Act of 1971 formally established the definition of a ‘cancer center’. Today there are a total of 72 National Cancer Institutes (NCI)-designated centers across the US delivering cancer care and conducting cutting-edge cancer research and clinical trials.^3^ All cancer centers are required to identify and serve a ‘catchment area’ defined as geographic areas from which the majority of its patients are drawn including the local area surrounding the facility.^4^

To best serve the needs of their catchment area, a clear understanding of cancer-specific data (including, epidemiological patterns, and risk factor prevalence), and community- and systemic- level drivers of health (healthcare access and social determinants) is critical for cancer centers to allocate resources, improve program efficiency, and maximize their reach and impact.

Beginning in 2012, the NCI has mandated that designated cancer centers identify and describe their catchment area and document ongoing research.^5^ However, the enormous volume of catchment area data, its complexity, the variety of data types, and the dispersion of data sources across federal and state agencies is a major challenge to synthesizing and relaying meaningful insights. An even bigger challenge is to effectively present these data to communities and a broad range of stakeholders (non-profit organizations, government agencies, researchers, cancer center leadership, clinical trial offices, payers, policymakers, and other groups - all with varying interests and expertise) to educate and empower them to take action. Data visualization (or DataViz) dashboards are important tools for gaining valuable insights from data.^6^ Data dashboards are widely used by industries and healthcare organizations for business intelligence as well as for tracking and monitoring their performance.^7, 8^ However, such dashboards remain untapped in the context of population health and are rarely developed with the end goal of community education and outreach. The main objective of this paper is to describe a systematic, stakeholder-driven, and theory-based approach for developing and evaluating catchment area data visualization tools for cancer centers. We present the approach, step-by-step, describing the personnel, material investments, data sources, and tools used to implement the approach.

## METHODS

Researchers at the Medical University of South Carolina (MUSC) Hollings Cancer Center (HCC) used a stakeholder-driven approach guided by a health communications framework. Cancer center researchers and community stakeholders worked collaboratively to develop an interactive catchment area data visualization dashboard, the South Carolina-Cancer Surveillance for Population Health Research and Outreach Tool (SC-SPOT), which was tested using a mixed-methods approach.

### Framework

The tool development process followed the Organize, Plan, Test, and Integrate (OPT-In) framework which is based on a variety of health communication theories and concepts **(Figure 1)**.^9^ The framework describes four core steps for sharing complex data or topics with a lay audience who may have a limited scientific background. A detailed description of the four steps of the framework and workflow is described below.

**FIGURE 1:**
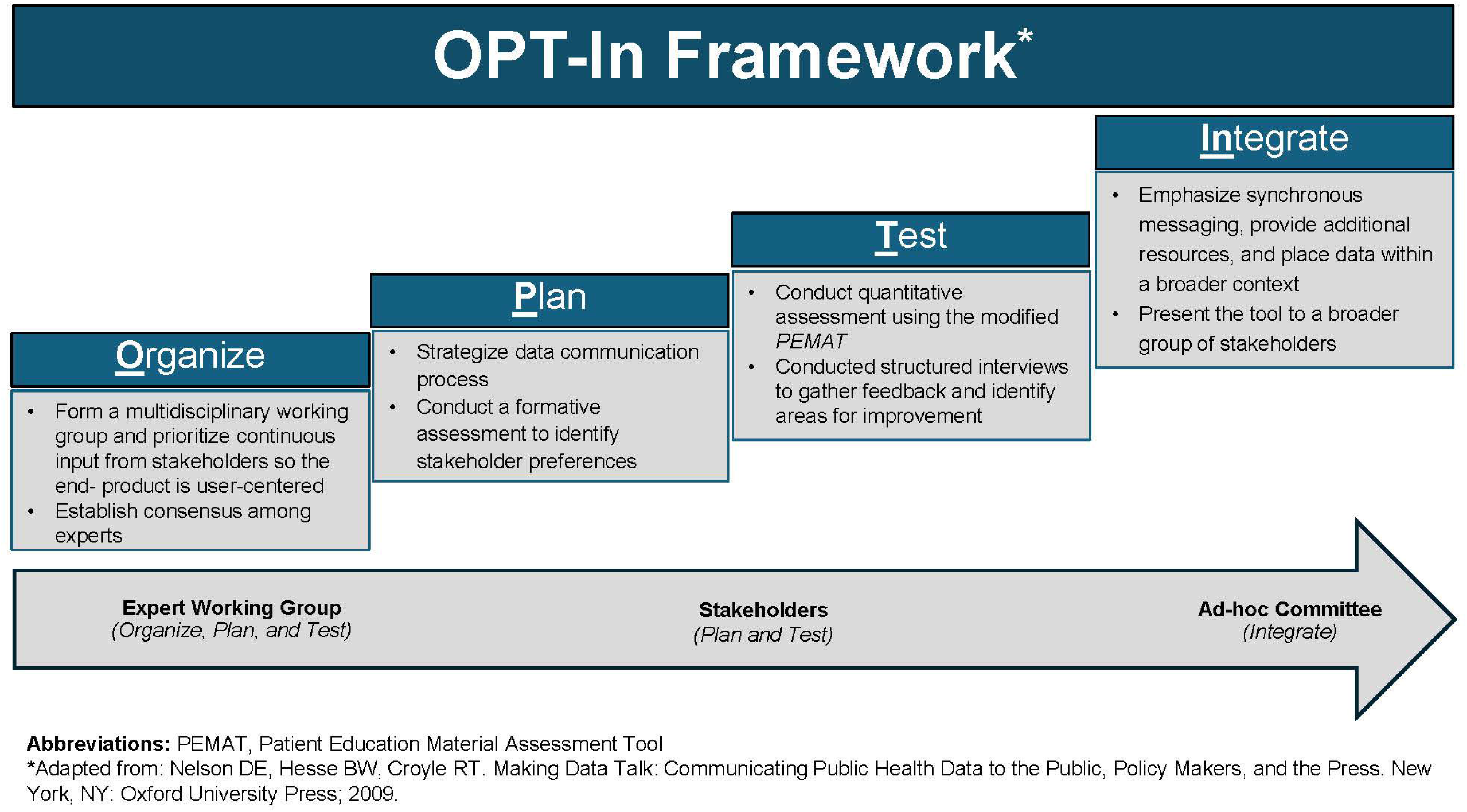
Framework for catchment area data visualization tool development. The figure illustrates the OPT-In framework and outlines key activities during each phase.

**STEP 1 Organize:** This step aims to develop a clear understanding of data and identification of end-users. A working group of multi-disciplinary experts including data analytics and visualization experts, epidemiologists, geospatial modelers, population health researchers, dissemination and implementation scientists, and community outreach and engagement experts is ideal. This group is responsible for the curation of all relevant data elements. Key sources of cancer-, sociodemographic-, census-, and healthcare-access-relevant data sources are identified from gold-standard data sources **(Table 1)**.

**TABLE 1:**
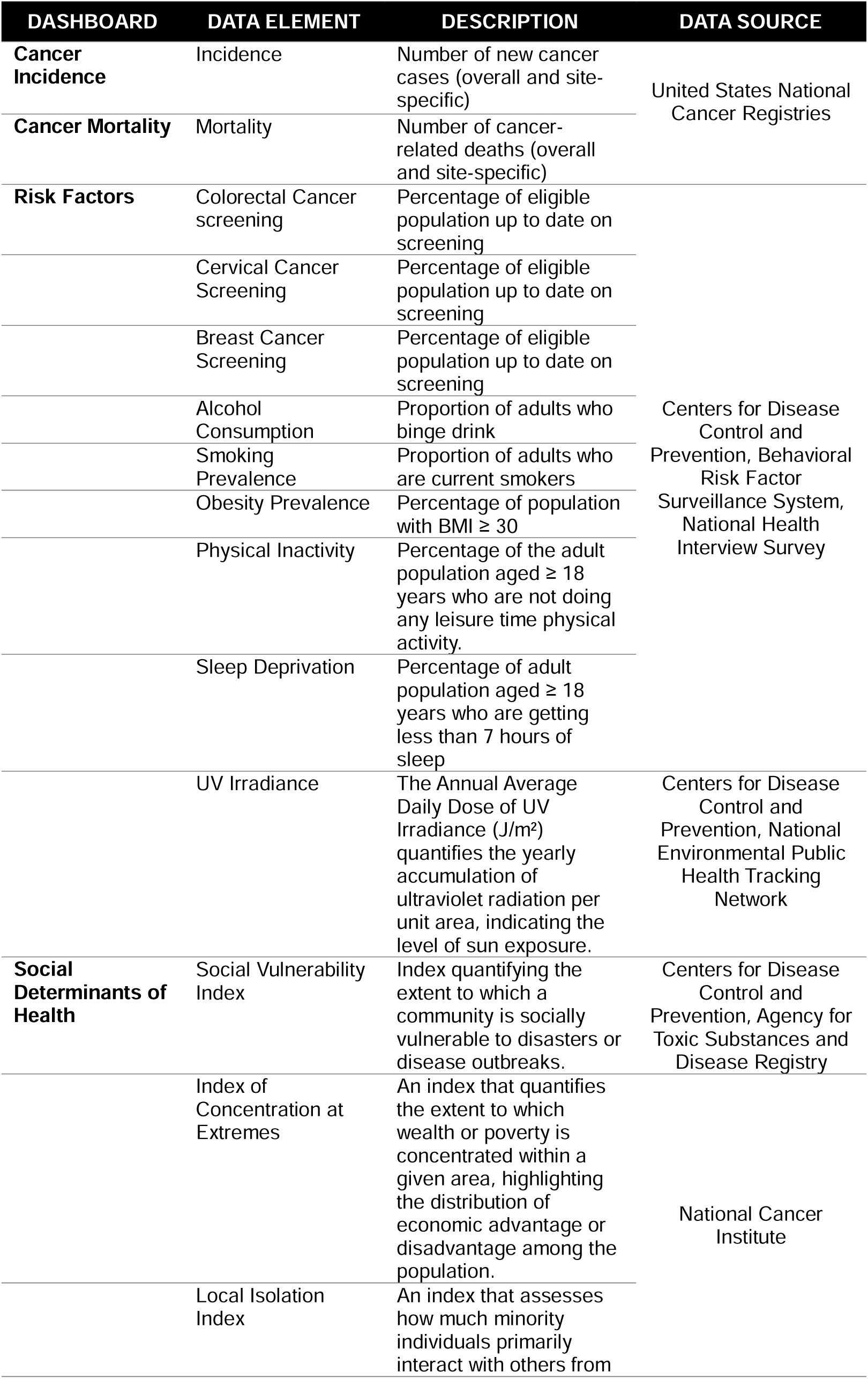
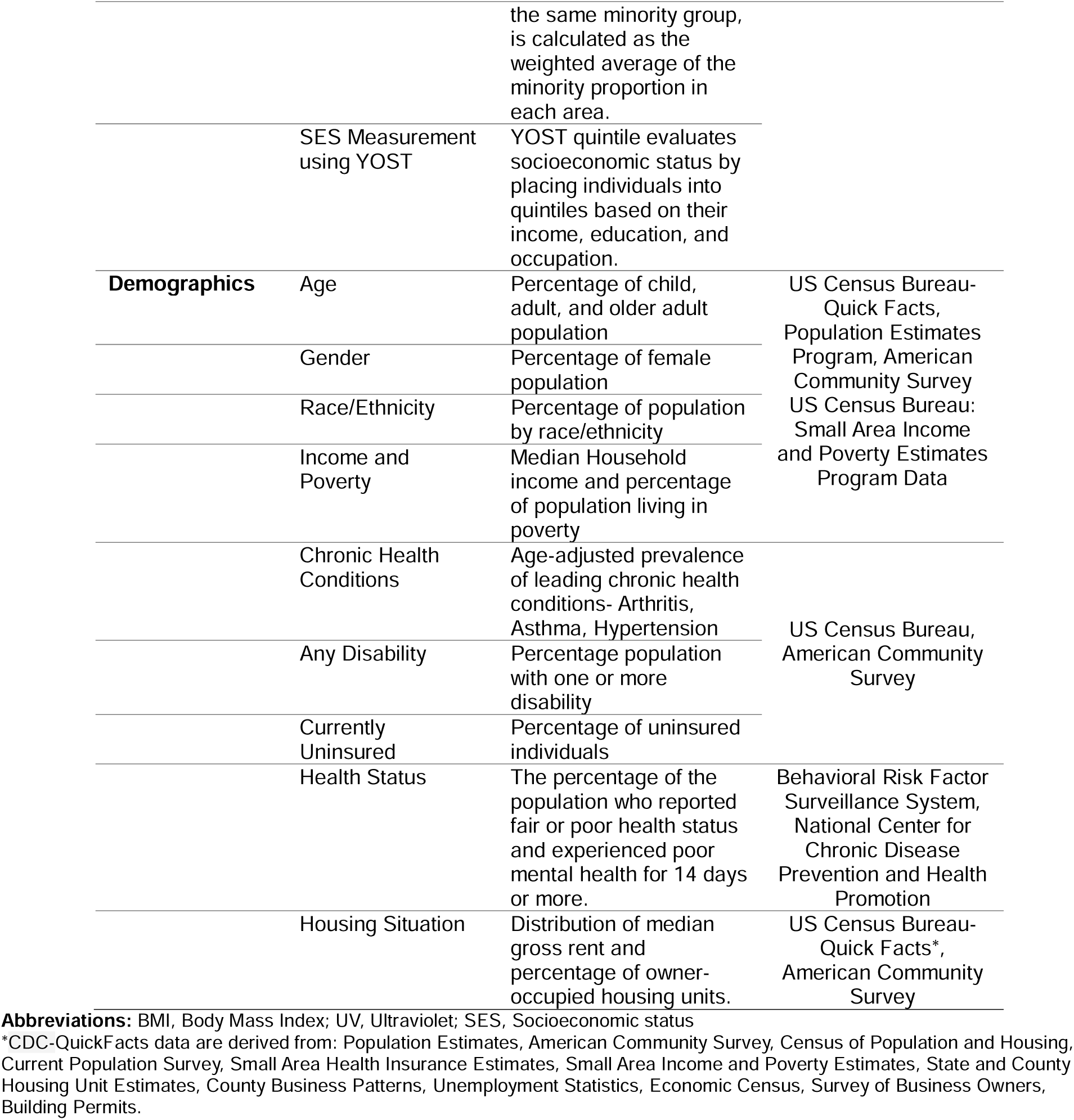
Data elements, description of elements, and their sources.

Making the tool user-centered is essential, therefore, continuously seeking input from the end-users (i.e., stakeholders) is integral to tool development. Given that the pool of end-users for catchment area data tools is broad, collaboration with diverse stakeholders is necessary. To develop SC-SPOT, we gathered feedback from our Community Advisory Board (CAB) including cancer patients, survivors, caregivers, representatives from non-profit organizations, payers, policymakers, government agencies, HCC clinicians and researchers, and SC residents.

**STEP 2 Plan:** The emphasis of this phase is the strategic presentation of data/information to the intended audience. This step is further broken down into four distinct processes: 1) Determining the purpose for communication, 2) Analyzing the audience(s), 3) Considering the context in which communication will occur, 4) Developing a preliminary product, and 5) Planning a strategy to reach audiences.

First, working group members should identify the communication goal central to the catchment area data tool. For instance, if the tool is primarily intended for dissemination and education across a range of end-users, then emphasis on understandability and actionability is most vital. If the tool is intended for researchers, administrators, and policymakers to quantify program or policy impact, the analytic capabilities of the tool are critical. Based on this central goal, all relevant audiences or end-users of the data should be identified. Third, contextual factors including the ease of access, self-navigation, and health literacy level of end-users should be carefully considered. A formative assessment can be conducted to identify relevant contextual factors as well as stakeholders’ priorities and preferences. Next, a prototype or minimum viable product must be developed as a ‘preliminary’ product with necessary functional capabilities (for example, data filtering, zooming (in and out of maps), search bars, and ready-to-download visuals) for testing. Finally, a mode of delivery must be identified which will maximize audience reach.

**STEP 3 Test:** This phase ascertains that the tool meets the communication goal. A mixed-methods approach for testing prototypes will generate both quantitative and qualitative endpoints. For instance, to test SC-SPOT, the HCC working group approached a set of naïve (uninvolved in the formative process in Step 2) 21 stakeholders, which included HCC CAB members, representatives from non-profit organizations, payers, policymakers, government agencies, HCC clinicians and researchers, and SC residents, with varying levels of familiarity with cancer-relevant data. For quantitative assessment, the use of an existing tool was deemed necessary. To our knowledge, there are currently no tools in the literature specifically designed to evaluate and test interactive data dashboards for stakeholder and community engagement. Therefore, the working group adopted and implemented a modified version of an education material assessment tool, known as the *Patient Education Material Assessment Tool (PEMAT),* developed by the Agency of Healthcare Research and Quality.^10^ The original *PEMAT* tool was developed to test educational material (print [P] or audiovisual [A/V]) on two parameters—1) Understandability (i.e., the education materials are *understandable* when users of diverse backgrounds and varying levels of health literacy can process and explain key messages.), and 2) Actionability (i.e., the education materials are *actionable* when consumers of diverse backgrounds and varying levels of health literacy can identify what they can do based on the information presented). *PEMAT* consists of a 24-item questionnaire with two possible responses agree (score=1) or disagree (score=0). We used a modified version of the *PEMAT* which included 20 of the 24 items that were relevant to data visualizations. The final score was calculated as a percentage (Total Points / Total Possible Points x 100), where a higher percentage indicates greater understandability and actionability.^11^ Next, to identify additional areas for improvement and feedback that were not captured through quantitative assessment, structured interviews with stakeholders were conducted. Interviews were transcribed and analyzed to identify areas for improvement.

**STEP 4 Integrate:** The final step of the framework focuses on integrating communication efforts and messages within a broader context of current scientific understanding. Emphasis on four aspects is recommended—1) synchronous messaging, 2) provision for additional resources, 3) providing a broader context to the data, and 4) directions for data usage. The emphasis on synchronous messaging is to ensure a consistent message across different channels. For example, the rates of new cancer cases presented on the tool should be consistent with cancer statistics presented on other pages on the website (including pages beyond the web tool). The provision for additional resources is to ensure users are provided or directed to resources beyond the tool. For example, dashboards presenting data on screening-detectable cancers can be accompanied by educational resources on cancer screening. Portraying information accurately, clearly, and in a useful manner is critical to providing a broader context to the data.^12^ Emphasis on minimizing the cognitive burden, ensuring accessibility, and using audience- tailored approaches will facilitate this aim. Finally, providing end-users guidance for correct interpretation and usage is imperative. At a minimum, a description of how data points were estimated and their inherent limitations can be specified.

## RESULTS

### Organization, Planning, and Prototype Development

The Working Group Members had recurring (bi-weekly) meetings to plan the process for communicating catchment area data. Thematic areas, data elements, and the composition and placement of the data visuals in the prototype were identified and finalized by the working group and the stakeholders. The group members acknowledged that communication of such complex data would require simplification through visuals and visual aids; therefore, a data visualization tool would be the most appropriate channel for relaying catchment area data. All relevant data elements were extracted from databases, transformed, and stored on a secured server. Data were converted into a format readable in the visualization software (Tableau^™^). All visualizations were developed and rendered in the *Tableau™* environment. The data visualization dashboards are embedded on the HCC public-facing website. Interactivity is maintained using the *Tableau Javascript* API. All interactive maps and graphics are color-blind friendly. All data visuals are downloadable in commonly used formats (Image, PowerPoint, and Excel) to facilitate dissemination. The working group recognized that sustainment of the tool beyond development will require resources for maintenance and regular updates as new data become available. The involvement of HCC leadership in the organization and planning process proved vital for gaining support to sustain the tool.

The visualization dashboards describe state- and county-level cancer incidence, mortality rates, calendar trends; burden (annual number of cases), the prevalence of risk factors, and social determinants of health. Interactive catchment area maps are color-coded based on data distribution to illustrate variation across geographic areas and to facilitate the identification of underperforming areas **(Figure 2)**. Data are presented at the most granular level available (county or zip-code tabulation area), with area-specific data displayed as users hover over the map. The dashboard also includes a search box where users can search for an area of interest. Calendar-year trends spanning over 20 years (measured as annual percentage changes) are illustrated using line graphs. Stratified data (race/ethnicity and sex-specific estimates) are presented as bar graphs. Drop-down menus allow the selection of optional data visualizations. For transparency regarding data sources, measures, and accuracy, an embedded document describing these methodological details is incorporated in webpages. The publicly accessible tool is available at https://hollingscancercenter.musc.edu/outreach/sc-spot-south-carolina-cancer-data-statistics.

**FIGURE 2:**
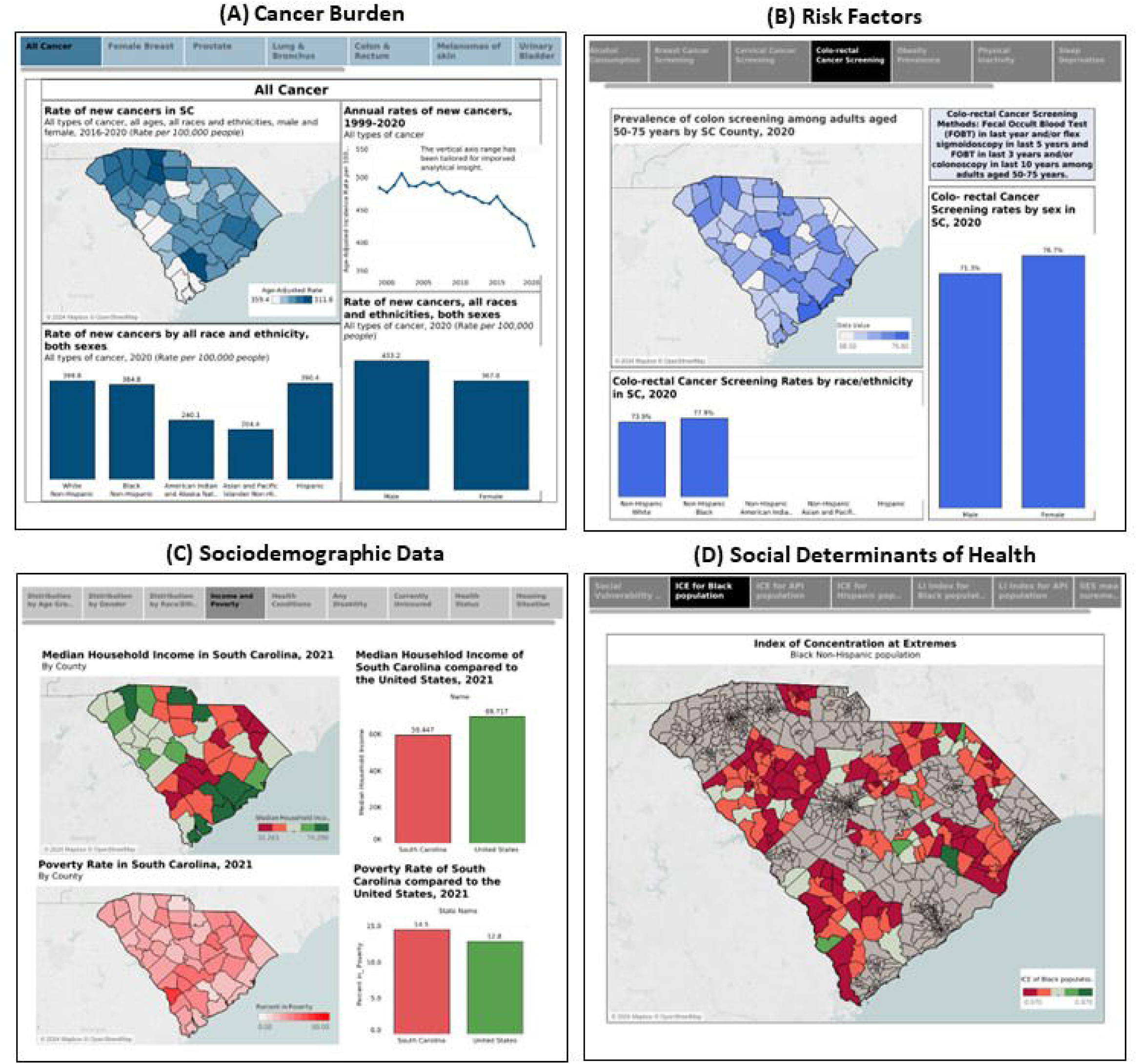
The Hollings Cancer Center catchment area data dashboard. The figure showcases key data visualization dashboards illustrating catchment area (i.e., the entire state of South Carolina) data for the Hollings Cancer Center: (A) Dashboard presents area-level variation, trends, and sociodemographic breakdown of cancer burden (i.e., incidence and mortality), (B) Dashboard illustrates the prevalence of risk factors and breakdown of prevalence by race and sex, (C) Dashboard captures key sociodemographic attributes of the catchment area population, and (D) Dashboard illustrates social determinants of health captured as composite measure illustrating vulnerability, segregation, isolation, and socioeconomic status.

### Testing

Quantitative and qualitative assessments were conducted with 21 naïve stakeholders (7 CAB members and other representatives, 10 clinicians and researchers, and 4 SC residents) selected through purposive sampling, each given a 5–8-minute overview of the tool and an opportunity to navigate the website. Data were collected using the modified *PEMAT and* open-ended responses to questions regarding the overall appeal and areas for improvement.

Results of the quantitative assessments of SC-SPOT, including a detailed breakdown of the modified *PEMAT* items and mean score, are presented in **Table 2**. The mean understandability score was 14.75 (±0.95; maximum possible 16 points). Mean actionability was 3.66 (±0.50; maximum possible 4 points).

**TABLE 2:** Understandability and actionability items adopted from the *PEMAT* and item-specific mean scores.

| **UNDERSTANDABILITY**<br>*Total Questions= 16 (Scoring: Agree=1; Disagree=0)* |  | **SCORE**<br>*Mean±SD* |
| --- | --- | --- |
| Content | This website is useful for understanding cancer data.<br>The website does not include information or content that is distracting | 2±0 |
| Word Choice & Style | The website uses common, everyday language.<br>When used, medical terms were easy to interpret | 1.75±0.50 |
| Use of Numbers | Numbers appearing on the website are clear and easy to understand.<br>The website expects the user to perform calculations.* | 2±0 |
| Organization | The website breaks or "chunks" data into short sections.<br>The website sections have informative headers.<br>The data presented on the website is confusing or not easy to understand.*<br>The website provides a summary of cancer data | 3.5±0.58 |
| Layout & Design | The website uses visuals like arrows, boxes, bullets, bold, larger font, and colors | 1±0 |
| Use of Visual Aids | The material uses graphs, so data is easily understood (e.g., maps, bar graphs).<br>The graphs make data interesting and reduce confusion.<br>The graphs have clear titles or captions.<br>The graphs are clear and uncluttered.<br>The website design is appealing (color, fonts, shapes) | 4.5±0.58 |
| **ACTIONABILITY**<br>*Total Questions= 4 (Scoring: Agree=1; Disagree=0)* |  |  |
| Information Seeking | I will use this website when I need cancer data | 1±0 |
| Information Sharing | I am likely to share this website with my family or friends if they need cancer data | 0.75±0.50 |
| Advocacy | Sharing data with the public and community is important.<br>The website will be useful to me and my community to advocate for better health or health care | 2±0 |

**Abbreviations:** PEMAT, Patient Education Material Assessment Tool; SD, Standard deviation
\*Responses were reverse coded Agree=0; Disagree=1

Open-ended responses suggest that the tool effectively communicates cancer data and has the potential for community education and advocacy. Additional areas of improvement that emerged were the use of a warm color palette, the inclusion of details regarding data sources, and providing benchmarks (i.e., presenting national-level data) for comparisons.

| **<u>EXAMPLES OF STAKEHOLDER FEEDBACK</u>** |
| --- |
| *<u>Comments on Overall Appeal</u>* |
| “This tool will be **useful** to show what issues we face as a state.”<br>“I think the website is very **informative**, I would **share** it with my colleagues.”<br>“I guess that the visualization tool looks **appealing** in the sense that it is **professional**.”<br>“This can be a great tool for inter-programmatic **engagement and collaboration**” |
| *<u>Comments on Design</u>* |
| “I think that if they utilize a lot more **different colors** maybe instead of just dark blue, it will look better.”<br>“You do need some **better colors**.”<br>“I would like to see more **earth-tone colors** used on the website.” |
| <u>Comments on Functionality</u> |
| "I want to look at social determinants of health (risk factors) **interlaid** with cancer incidence (outcomes)"<br>"Having information by house and senate districts would be ideal for **advocacy.**"<br>"I would like to see how our state is doing **compared to others.**" |
| <u>Areas for Improvement</u> |
| "You should say where the data came from (**data source**)"<br>"I noticed there were **no percentage bars** for cervical cancer screening for American Indian and Hispanic women, does that mean no one underwent screening in those groups?" |

### Integration

Currently, in progress, the HCC working group plans to address the four recommended areas for integration—synchronous messaging, provision for additional resources, providing a broader context to the data, and directions for data usage—through an ad-hoc committee. The ad-hoc committee is responsible for critically evaluating content on the website, including webpages outside of the dashboard interface, to ensure that only high-quality evidence is cited and that evidence-based strategies for cancer prevention, treatment, and survivorship are emphasized. The committee is responsible for ensuring that information shared on these topics is consistent across the web pages to maintain synchronicity in messaging. Additional resources will be highlighted across the HCC’s web pages to navigate users beyond the visualization tool. These will include 1) educational material on cancer screening, tobacco cessation, and cancer survivorship, 2) options for scheduling mobile cancer screening and vaccinations, 3) identifying oncologists, 4) resources for financial counseling, 5) genetic counseling information, and 6) information for clinical trial participation. Additionally, two experts from the working group responsible for demonstrating the tool will provide directions for data usage during presentations to state agencies, non-profit organizations, and policymakers. Dissemination activities and analytical data (as of June 2024) on users are summarized in **Table 3**.

**TABLE 3:** SC-SPOT dissemination activities and user analytics (as of June 2024).

| **DISSEMINATION ACTIVITIES** |  |  |  |  |  |  |
| --- | --- | --- | --- | --- | --- | --- |
| *Invited Presentations (including external and internal)* |  |  |  |  |  |  |
| South Carolina Cancer Alliance<br>Office of Senator Lindsay Graham<br>BlueCross BlueShield of South Carolina<br>South Carolina Department of Health and Environmental Control Office of Cancer Control<br>American Medical Informatics Association<br>Catchment Area Data Conference<br>MUSC Social Determinants Of Health working group<br>MUSC Enterprise Chief Digital Transformation Office<br>MUSC Population Health Leadership Committee<br>Hollings Cancer Center Executive Advisory Board<br>Hollings Cancer Center Cancer Control Program<br>Hollings Cancer Center Clinical Trials Office |  |  |  |  |  |  |
| *News outlets/ Newsletters* |  |  |  |  |  |  |
| WSBC News<br>Fox News 24<br>Post and Courier<br>Health IT Analytics<br>AMIA Informatics Smartbrief |  |  |  |  |  |  |
| **USER ANALYTICS** *(Data is for the period of 3/15/24-6/11/24)* |  |  |  |  |  |  |
| PAGE | VIEWS | ACTIVE USERS | SESSIONS | ENGAGED SESSIONS | ENGAGEMENT RATE* | AVERAGE SESSION DURATION |
| Landing Page | 779 | 439 | 633 | 348 | 54.91% | 1:32 |
| New Cancer Cases | 278 | 167 | 225 | 192 | 85.33% | 1:52 |
| Social determinants | 196 | 139 | 177 | 148 | 83.62% | 1:39 |
| Cancer-Related Deaths | 118 | 92 | 113 | 99 | 87.61% | 1:48 |
| Cancer Risk Factors | 107 | 78 | 97 | 86 | 88.66% | 2:42 |
| Demographics | 73 | 56 | 75 | 64 | 85.33% | 1:27 |
| Health Care Access | 57 | 44 | 51 | 44 | 86.27% | 1:54 |
| **TOTAL** | **1608** | **541** | **834** | **448** | **86.14%** | **1:51** |

\*Total engagement rate reflects engagement with dashboards (i.e., excludes landing page).

## DISCUSSION

With the increased importance of data-driven decision-making in medicine, community engagement, and population health, there is a growing demand for tools that can efficiently and effectively present complex data in an easy-to-digest format.^6^ Identification and characterization of the cancer burden and contributing factors in catchment areas is a central goal across all NCI Designated Cancer Centers.^5^ Remarkably, the need for catchment area data is intertwined with the broader goal of improving the cancer center’s community outreach and engagement. Thus, catchment area data necessarily need to be accessible and useful to a wide audience (communities, non-profit organizations, government agencies, researchers, clinical trial offices, payers, policymakers, and other groups) with varying interests and expertise. Understanding and incorporating the preferences and needs of these broad groups of end-users when developing catchment area data resources is imperative.^13^ By actively engaging stakeholders from its inception, we were able to generate an innovative catchment area data resource that conforms with the needs of heterogeneous end-users. Furthermore, through validated tools and qualitative data collection, we were also able to objectively quantify the understandability and actionability of the tool and garner subjective feedback for improvisation.

Collaboration between population health researchers, basic scientists, clinicians, information technology experts, and stakeholders is foundational for developing catchment area data resources. For instance, the expertise of population health researchers and clinicians was necessary to gather credible sources of data and identify metrics that accurately capture burden and risk factors, and to create a blueprint of the data visualization dashboard. Identifying and procuring cutting-edge information technology infrastructure (including storage, data architecture, web design, and interactivity between the back end and public-facing components) was essential for the efficient execution of the dashboards and ensuring a seamless end-user experience. Achieving the delicate balance between information load and comprehension integral to human-computer interaction would have been impossible to achieve without input from key stakeholders. Each group brought unique strengths and perspectives which led to a robust, easily accessible, and shareable user-centered catchment area data resource. Each end-user can leverage the information to further strengthen their mission and/or to make collective data-driven decisions.^14^

Based on spatial patterns observed in the data, community outreach, and engagement staff can target geographic areas in need of intervention.^5^ Such activities could be tailored further by understanding the sociodemographic composition of the target communities. Towards these efforts, the catchment area data visualization tool could be used as a ‘primer’ for educating members of these communities about cancer in their communities and to build trust and confidence for engagement activities. Overall, within the context of community outreach, catchment area data tools could substitute ‘needs assessment’ reports,^15^ a priori often mandatory to justify effort allocation and guide the development of community action plans.

From the research standpoint, basic and population health scientists could leverage information from the tool to describe the cancer burden and risk factor contribution to create a case for research investments in understanding the basic biology and mechanisms, development of new therapeutics, translational science, and developing, testing, and implementing medical innovations and evidence-based interventions.

Recent federal mandates to capture and address social determinants of health emphasize the need for healthcare providers and clinical staff to be aware and knowledgeable on relevant metrics such as the built environment, healthcare access, socioeconomic standing, education, and social factors (segregation, isolation, discrimination, and lack of social support) of their catchment areas.^16^ Furthermore, at the health system level, catchment area data tools can also be critical for executive leadership in creating a business case for expansion in medically underserved areas or for planning and strategizing new programs and/or initiatives across the healthcare enterprise.

Finally, the prospect of catchment area data visualization tools contributing to the development and implementation of data-driven policies is promising. However, to achieve this end goal, it may be necessary to first identify and define an issue and then present it to policymakers in a compelling manner backed with data. Next, it will also be critical to invigorate advocacy at the grassroots level to build and strengthen coalitions between stakeholders. Through visualizations, catchment area data tools can also present compelling data stories which can be useful for educating and communicating key takeaways to both policymakers and key stakeholders such as non-profit organizations, coalitions, patient advocacy groups, and community members.

Limitations of the data visualization tool should also be carefully considered. The data presented on the visualization dashboard were collected by multiple federal- or state-level agencies, employing specific data collection techniques (surveillance systems, telephone surveys, home interviews, etc.). Each database has varying degrees of sensitivity, specificity, and limitations inherent to the data collection process. For example, data from surveys are prone to recall bias and social desirability bias. Mortality ascertainment is not possible for all cancer cases in cancer registries. Strict patient confidentiality laws relevant to patient identifiers (including area of residence) may limit the granularity of data visualization. Due to the disintegrated nature of healthcare in the US and the lack of linkages across databases, having complete information on individuals is not always possible. For instance, information on HIV/AIDS status, a major clinical risk factor for certain cancers, is not available in cancer registries. Despite their limitations, these data sources provide valid and reliable estimates relevant to the US population with opportunities for data cross-linkage (e.g., HIV/AIDS registry linkage with cancer registry) when necessary.

In conclusion, cancer centers in the US are key infrastructures for preventing, treating, and eliminating cancers from US communities. In addition to providing cancer care, cancer centers also shoulder the responsibility of community education, outreach, and engagement, advancing research, and workforce training. The SC-SPOT tool created by the Hollings Cancer Center exemplifies the importance of stakeholder engagement for creating catchment area data resources. Taking this approach ensures that the end-users will have a seamless interactivity experience, the cancer information is relayed effectively, and stakeholders gain actionable insights. Generating a catchment area data resource, such as the SC-SPOT, is a necessary first step. Making a measurable impact will require time, resources, and a long-term plan for continuous outreach and advocacy by cancer centers.

## Data Availability

Data used in the study are publicly available on the SEER, Census, and CDC website.

https://seer.cancer.gov/

https://www.cdc.gov/index.html

https://www.census.gov/data.html

## Acknowledgments

We sincerely thank all stakeholders for their engagement and participation. We would also like to thank Dan Rinder and William Morgenweck for providing development support and Alexis Nuzzo, MPH for her help with proofreading the content.

